# Magnitude and Factors Associated with Hypertension among Conflict-affected Adults in Northern Ethiopia: A Community-Based Cross-Sectional Study

**DOI:** 10.64898/2026.09.03.26362189

**Authors:** Andnet Tadesse Wete, Simon P.S. Kibira, Thomas Buyinza, Dathan M. Byonanebye

**Author notes:** Corresponding author: Andnet Tadesse, Department of Community Health and Behavioral Sciences, of Makerere University, Kampala, Uganda.

## Abstract

**Background:** Hypertension is a growing public health problem worldwide; however, evidence of the burden and determinants in populations affected by conflict and displacement remains limited. This study aimed to determine the magnitude of hypertension and associated factors among conflict-affected adults.

**Methods:** We conducted a community based, cross-sectional study in September and October 2025, among 839 adults aged 25-65 years who were internally displaced, returnees or conflict survivors. Socio-demographics, wealth status, behavioral characteristics, mental health and conflict related experiences data were collected using interview-administered questionnaires following standardized blood pressure and anthropometric measurements. Modified Poisson regression with robust variance was used to estimated crude and adjusted prevalence ratios. Factors with p < 0.05 were considered statistically significant.

**Results:** Overall, 839 were enrolled, 467 (55.7%) were female, the median age was 41 (IQR: 35-46) years, and 264 (31.5%) had hypertension. Factors associated with hypertension were age >44 years, bereavement (aPR=1.89, 95%CI: 1.60-2.25) experiencing major property loss (aPR=1.34, 95%CI: 1.01-1.77), psychological stress (aPR=1.99, 95%CI: 1.57-2.52), lower physical activity (aPR=2.02, 95CI: 1.24-3.30) and processed food consumption (aPR=1.36, 95%CI: 1.17-1.59).

**Conclusion:** The burden of hypertension among conflict–affected populations in Ethiopia is high. Hypertension was significantly associated with several socio-demographic, behavioral, psychosocial, and biological factors. These findings highlight the need for integrated, context-specific interventions that combine cardiovascular care with mental health support, social protection, and strengthened primary health systems in conflict-affected settings.

## Introduction

Hypertension is a major public health problem that has not been fully addressed among populations affected by conflict [1]. Globally, 117.3 million people had been forced to flee their homes due to conflict and human rights violations [2], where disruption of healthcare systems, psychological stress, and displacement may increase the burden of hypertension. Evidence indicates that casualties arising from conflict-related incidents often exceed the rates of morbidity and mortality typically observed in areas unaffected by conflict, impacting both military personnel and civilians [1, 3].

According to the United Nations High Commissioner for Refugees (UNHCR) aggregated estimate of Sub-Saharan Africa (SSA) and the Middle East and North Africa (MEND), 61 million people are experiencing humanitarian crises either being displaced or refugees [4]. In 2024, there were more than 45 million people displaced or refugees in Africa [5]. In Ethiopia, approximately 2,816,890 people were forcibly displaced in 2023 [6] and healthcare system disruptions were among major challenges of the community [7]. However, the prevalence of hypertension was not assessed well in conflict-affected settings.

In the general population studies in Africa, hypertension is known as a leading cause of premature deaths and the estimated prevalence is around 27.2% [8]; there is strong evidence linking socio-demographic and several modifiable risk factors, such as unhealthy foods, sedentary lifestyles, being overweight or obese, cigarette use, and alcohol abuse, with an increased risk of hypertension [1, 8–9]. However, psychosocial variables were rarely included in most of hypertension predictors’ targeted studies and even the interaction between mentioned factors and hypertension in conflict settings is yet unclear.

Similarly, SSA which experiences one of the greatest refugee crises, the burden and drivers of hypertension among conflict-affected populations remain unclear. Therefore, we integrated the concept of bio-psychosocial model (BPM) and the social determinants of health (SDH) to holistically examine and unpack biological, psychological, and socio-structural factors [10, 11], in addition to determining the prevalence of hypertension in conflict-affected people of Amhara region, Ethiopia.

## Materials and Methods

### Study area and period

The study was conducted between September and October 2025 in the Amhara region of Ethiopia, which had experienced prolonged armed conflict from Nov. 2020 to Nov. 2022 resulting in large-scale population displacement [12].

### Study design, and population

A community-based cross-sectional study was conducted using kebele-level sampling frames. The study population comprised all civil conflict–affected individuals aged 25–65 years in the Amhara region. Eligible participants were permanent residents (≥6 months) of Amhara conflict-affected Zones, Woredas and Kebeles who were identified as IDPs, returnees, or crisis survivors. Those who did not provide written consent, pregnant women, the severely ill, and individuals with known mental health disorders were excluded.

### Sample size determination and sampling procedures

The prevalence of hypertension in conflict settings is unknown and 50% prevalence was assumed to generate the highest sample size. Assuming, 80% power, 95% confidence level and 5% margin of error. The initial sample size was calculated to be 384. To account for a potential 10% non-response rate, the sample size was increased to 427. The sample size was inflated by a design effect of 2 [13] to account for multistate sampling, resulting in the final sample size of 854 participants.

A multistage sampling technique was employed. Initially, two zones and one city administration were randomly selected from the seven zones and one city administration affected by the conflict [14]. Subsequently, districts within the selected administrative units were stratified into urban, semi-urban, and rural strata, from which one district was randomly selected per stratum. In the next stage, one kebele (the smallest administrative unit of the local government) was randomly selected from each district. Finally, households were selected from an updated sampling frame of each kebele based on its population size using simple random sampling via the lottery method until a total of 854 households were obtained. In each selected household, one eligible participant was randomly chosen for interview.

## Measurements and definitions

### Study variables

outcome (Hypertension); independent variables include socio-demographic variables, behavioral variables (alcohol intake, cigarette smoking, kihat chewing, number of substances used following the conflict and dietary habits), psychosocial variables (Chronic Psychological stress, bereavement and major property loss due to conflict), clinical and anthropometric variables (blood pressure, weight and height). Individuals were considered to have hypertension if they had a systolic blood pressure ≥ 140 mmHg and/or a diastolic blood pressure ≥ 90 mmHg taken at least five minutes apart or if they were receiving antihypertensive medications during the time of data collection in the study area, Amhara region Ethiopia.

Hypertension control was defined as the achievement of sustained blood pressure readings consistently below 140/90 mmHg,using standard techniques, in patients diagnosed with hypertension. Body mass index: underweight (<18.5 kg/m^2^), normal (18.5–24.9 kg/m^2^), overweight (25.0–29.9 kg/m^2^) and obese (>30.0 kg/m^2^). Substances used following the conflict stress in the setting include: tobacco, alcohol, khat, cannabis, shisha and local stimulant.

### Data collection instruments and procedures

An interviewer administered questionnaire was used after 5% tested for data collection; it was adapted from the WHO ‘STEP wise approach tool for NCD surveillance [15] and included most of the questions in core version and some of the questions in expanded version. Wealth Index (WI) was assessed through durable assets ownership, housing characteristics, water and sanitation, utilities and energy, as well as agricultural/rural assets; and also number of substances used, locally relevant items like Khat chewing [16], fruit and vegetable consumption, Global Physical Activity Questionnaire (GPAQ) to measure physical activity level [17], conflict related questions like bereavement and major property loss were incorporated.

Anthropometric and blood pressure measurements were conducted following WHO standard procedures [15]. Participants rested at least for 5 minute prior to 3 blood pressure measurements were taken at 1–2-minute intervals using an automated digital monitor with their feet flat on the floor and the arm supported at heart level; whereas, those with recent intake of heavy meal, exercise, smoking or caffeine intake were delayed for at least 30 minutes before measurement and instructed to rest quietly for 5 minutes in a seated position. If consecutive readings differed by >10 mmHg systolic or >6 mmHg diastolic, an additional measurement was obtained after a further 5-minute rest. The average of the second and third readings was used for analysis.

Further, psychological and shared community stress was assessed using adapted versions of the Perceived Stress Scale (PSS-10) [18, 19]; both tools, α = .766 and α = .721 respectively, demonstrated acceptable preliminary internal consistency following pilot testing.

### Quality assurance

The training conducted for data collectors ensured a clear understanding of the procedures and study protocols. Pre-tests done prior to the actual data collection helped refine the tools and identify potential challenges. Supervision performed during the data collection process ensured adherence to the study guidelines and maintained data accuracy, while standardized procedures used throughout the activity promoted consistency, reliability, and quality of the collected data.

### Data management and statistical analysis

Data were entered and cleaned using Epi Info version 3.5.4 and then exported to SPSS. Further data cleaning and statistical analyses were conducted using SPSS version 27. Modified Poisson regression was employed as the primary analytical approach to estimate prevalence ratios.

Principal component analysis (PCA) was performed to construct a household wealth index; the first principal component, which explained 42.2% of the total variance, was retained to classify households into wealth quintiles for subsequent analyses.

The internal consistency analysis was conducted for both psychological and shared community stress tools using the main dataset and demonstrated: α = .872 and α = .858 respectively.

Descriptive analyses were conducted to summarize socio-demographic, conflict–related, behavioral, and comorbidity characteristics. Categorical variables were presented as frequencies and percentages, while continuous variables, such as blood pressure, were summarized using means and standard deviations.

Bivariate analyses were performed using modified Poisson regression with robust variance estimation to calculate crude prevalence ratios (cPR) and 95% confidence intervals for the association between each independent variable and hypertension. Variables with a p-value < 0.25, as well as those deemed theoretically relevant or previously established as associated with hypertension, were considered for inclusion in the multivariable model [20].

Multivariable analysis was then conducted using modified Poisson regression to identify factors independently associated with hypertension. Adjusted prevalence ratios (aPR) with 95% confidence intervals were reported after assessing multicollinearity using variance inflation factors and tolerance tests [21, 22]. Variables with a p-value < 0.05 were considered statistically significant and reported as factors associated with hypertension among civil crisis–affected populations.

### Ethics approval and consent to participate

Ethical approval and clearance for this study was initially obtained from the Makerere University School of Public Health Research Ethics Committee (SPH-2025-886), Uganda and then from the Amhara Regional Research Ethical Review Board (ARRERB Ref. no: <u>NoH/R/De/Di/07/114</u>),

Ethiopia. The overall participation was voluntary and each study participant was informed about the research objectives, methods and techniques in detail and written informed consent was obtained from all participants prior to their inclusion in the study.

## Results

We obtained a 98.2% response rate and overall, 839 conflict affected individuals were included in the analysis, 467(55.7%) were females and 372 (44.3%) were males (Table 1). Their median age was 41 years (IQR: 35-46). The highest number, 345(41.1%) of study participants were crisis survivors, while 326(38.9%) were returnees, and 168(20.0%) were IDPs. Majority, 367(43.7%) resided in urban areas, 730(87.0%) were married, and 470(56.0%) had completed secondary education.

**Table 1.** The characteristics of study participants stratified by crisis-driven status (n=839)

| Variable | Category | IDP (n=168) | Returnee (n=326) | Crisis survivor (n=345) | Total (n=839) |
| --- | --- | --- | --- | --- | --- |
| **Sex of study participants** |  |  |  |  |  |
|  | Female | 81 (48.2%) | 189 (58.0%) | 197 (57.1%) | 467 (55.7%) |
|  | Male | 87 (51.8%) | 137 (42.0%) | 148 (42.9%) | 372 (44.3%) |
| **Age categories (years)** |  |  |  |  |  |
|  | 25-34 | 20 (11.9%) | 97 (29.8%) | 78 (22.6%) | 195 (23.2%) |
|  | 35-44 | 76 (45.2%) | 138 (42.3%) | 143 (41.5%) | 357 (42.6%) |
|  | 45-54 | 50 (29.8%) | 76 (23.3%) | 88 (25.5%) | 214 (25.5%) |
|  | 55-65 | 22 (13.1%) | 15 (4.6%) | 36 (10.4%) | 73 (8.7%) |
| **Location of residence** |  |  |  |  |  |
|  | Urban | 60 (35.7%) | 145 (44.5%) | 162 (47.0%) | 367 (43.7%) |
|  | Semi-urban | 45 (26.8%) | 71 (21.8%) | 70 (20.2%) | 186 (22.2%) |
|  | Rural | 63 (37.5%) | 110 (33.7%) | 113 (32.8%) | 286 (34.1%) |
| **Marital status** |  |  |  |  |  |
|  | Not married | 9 (5.4%) | 55 (16.9%) | 45 (13.0%) | 109 (13.0%) |
|  | Married | 159 (94.6%) | 271 (83.1%) | 300 (87.0%) | 730 (87.0%) |
| **Educational status** |  |  |  |  |  |
|  | Primary education/less | 36 (21.4%) | 45 (13.8%) | 48 (13.9%) | 129 (15.4%) |
|  | Secondary education | 120 (71.4%) | 165 (50.6%) | 185 (53.6%) | 470 (56.0%) |
|  | Higher education | 12 (7.2%) | 116 (35.6%) | 112 (32.5%) | 240 (28.6%) |
| **Occupational status** |  |  |  |  |  |
|  | Employed | 118 (70.2%) | 259 (79.4%) | 273 (79.1%) | 650 (77.5%) |
|  | Unemployed | 50 (29.8%) | 67 (20.6%) | 72 (20.9%) | 189 (22.5%) |
| **Bereavement/lost family member/s** |  |  |  |  |  |
|  | Yes | 71 (42.3%) | 90 (27.6%) | 118 (34.2%) | 279 (33.3%) |
|  | No | 97 (57.7%) | 236 (72.4%) | 227 (65.8%) | 560 (66.7%) |
| **Experienced major property loss** |  |  |  |  |  |
|  | Yes | 166 (98.8%) | 294 (90.2%) | 307 (89.0%) | 767 (91.4%) |
|  | No | 2 (1.2%) | 32 (9.8%) | 38 (11.0%) | 72 (8.6%) |
| **Suffered physical injury/any harm** |  |  |  |  |  |
|  | Yes | 87 (51.8%) | 159 (48.8%) | 193 (55.9%) | 439 (52.3%) |
|  | No | 81 (48.2%) | 167 (51.2%) | 152 (44.1%) | 400 (46.7%) |
| **Substance use** |  |  |  |  |  |
|  | Yes | 79 (47.0%) | 60 (18.4%) | 96 (27.8%) | 235 (28.0%) |
|  | No | 89 (53.0%) | 266 (81.6%) | 249 (72.2%) | 604 (72.0%) |
| **Chronic Psychological Stress (CPS)** |  |  |  |  |  |
|  | High | 104 (61.9%) | 199 (61.0%) | 199 (57.7%) | 502 (59.8%) |
|  | Moderate | 64 (38.1%) | 127 (39.0%) | 146 (42.3%) | 337 (40.2%) |
| **Shared/Common Community Stress (CCS)** |  |  |  |  |  |
|  | High | 133 (79.2%) | 67 (20.6%) | 68 (19.7%) | 268 (31.9%) |
|  | Moderate | 35 (20.8%) | 259 (79.4%) | 277 (80.3%) | 571 (68.1%) |
| **Tobacco smoking** |  |  |  |  |  |
|  | Yes | 16 (9.5%) | 16 (4.9%) | 27 (7.8%) | 59 (7.0%) |
|  | No | 152 (90.5%) | 310 (95.1%) | 318 (92.2%) | 780 (93.0%) |
| **Exposure to smoking** |  |  |  |  |  |
|  | Yes | 6 (3.6%) | 6 (1.8%) | 14 (4.1%) | 26 (3.1%) |
|  | No | 162 (96.4%) | 320 (98.2%) | 331 (95.9%) | 813 (96.9%) |

**Table 1. (Continued)**
| Variable | Category | IDP (n=168) | Returnee (n=326) | Crisis survivor (n=345) | Total (n=839) |
| --- | --- | --- | --- | --- | --- |
| **Consuming alcohol** |  |  |  |  |  |
|  | Yes | 46 (27.4%) | 32 (9.8%) | 59 (17.1%) | 137 (16.3%) |
|  | No | 122 (72.6%) | 294 (90.2%) | 286 (82.9%) | 702 (83.7%) |
| **Khat chewing** |  |  |  |  |  |
|  | Yes | 38 (22.6%) | 34 (10.4%) | 55 (15.9%) | 127 (15.1%) |
|  | No | 130 (77.4%) | 292 (89.6%) | 290 (84.1%) | 712 (84.9%) |
| **Limit processed food consumption** |  |  |  |  |  |
|  | Yes | 113 (67.3%) | 211 (64.7%) | 212 (61.4%) | 536 (63.9%) |
|  | No | 55 (32.7%) | 115 (35.3%) | 133 (38.6%) | 303 (36.1%) |
| **Limit salty sauce utilization** |  |  |  |  |  |
|  | Yes | 87 (51.8%) | 192 (58.9%) | 206 (59.7%) | 485 (57.8%) |
|  | No | 81 (48.2%) | 134 (41.1%) | 139 (40.3%) | 354 (42.2%) |
| **Physical activity (PA)** |  |  |  |  |  |
|  | Low | 82 (48.8%) | 69 (21.2%) | 95 (27.5%) | 246 (28.3%) |
|  | Moderate | 81 (48.2%) | 228 (69.9%) | 206 (59.7%) | 515 (61.4%) |
|  | High | 5 (3.0%) | 29 (8.9%) | 44 (12.8%) | 78 (9.3%) |
| **Have diabetes** |  |  |  |  |  |
|  | Yes | 19 (11.3%) | 24 (7.4%) | 19 (5.5%) | 62 (7.4%) |
|  | No | 149 (88.7%) | 302 (92.6%) | 326 (94.5%) | 777 (92.6%) |
| **Family history of hypertension** |  |  |  |  |  |
|  | Yes | 26 (15.5%) | 33 (10.1%) | 45 (13.0%) | 104 (12.4%) |
|  | No | 142 (84.5%) | 293 (89.9%) | 300 (87.0%) | 735 (87.6%) |
| **Body Mass Index (BMI) category** |  |  |  |  |  |
|  | Normal | 65 (38.7%) | 219 (67.2%) | 212 (61.5%) | 496 (59.1%) |
|  | Overweight | 69 (41.1%) | 87 (26.7%) | 88 (25.5%) | 244 (29.1%) |
|  | Obese | 34 (20.2%) | 20 (6.1%) | 45 (13.0%) | 99 (11.8%) |

Concerning bereavement, 279 (33.3%) study participants reported loss of family member/s, 767(91.4%) reported major property loss and 337(40.2%) had moderate level of chronic psychological stress. The study result revealed that 59(7.0%) of study participants reported current tobacco use; 137(16.3%) reported alcohol consumption within the past 30 days, 235(28%) reported substance use and 246(29.3%) of participants had low physical activity level.

Among 839 study participants, 62 (7.4%) had diabetes mellitus, while 104 (12.4%) reported a family history of hypertension. The median body mass index (BMI) was 23.6kg/m² (IQR: 22.86-26.14), the median systolic and systolic blood pressure were 120mmhg (IQR: 116-130) and 72mmhg (IQR: 67-80) respectively.

### Hypertension prevalence and control

The overall prevalence of hypertension in this population was 31.5%. (95% CI: 28.3-34.6%). When stratified by crisis-driven status, 48.8% of IDPs, 25.2% of returnees and 29.0% of crisis survivors were hypertensive (**Figure 1**).

**Figure 1:**
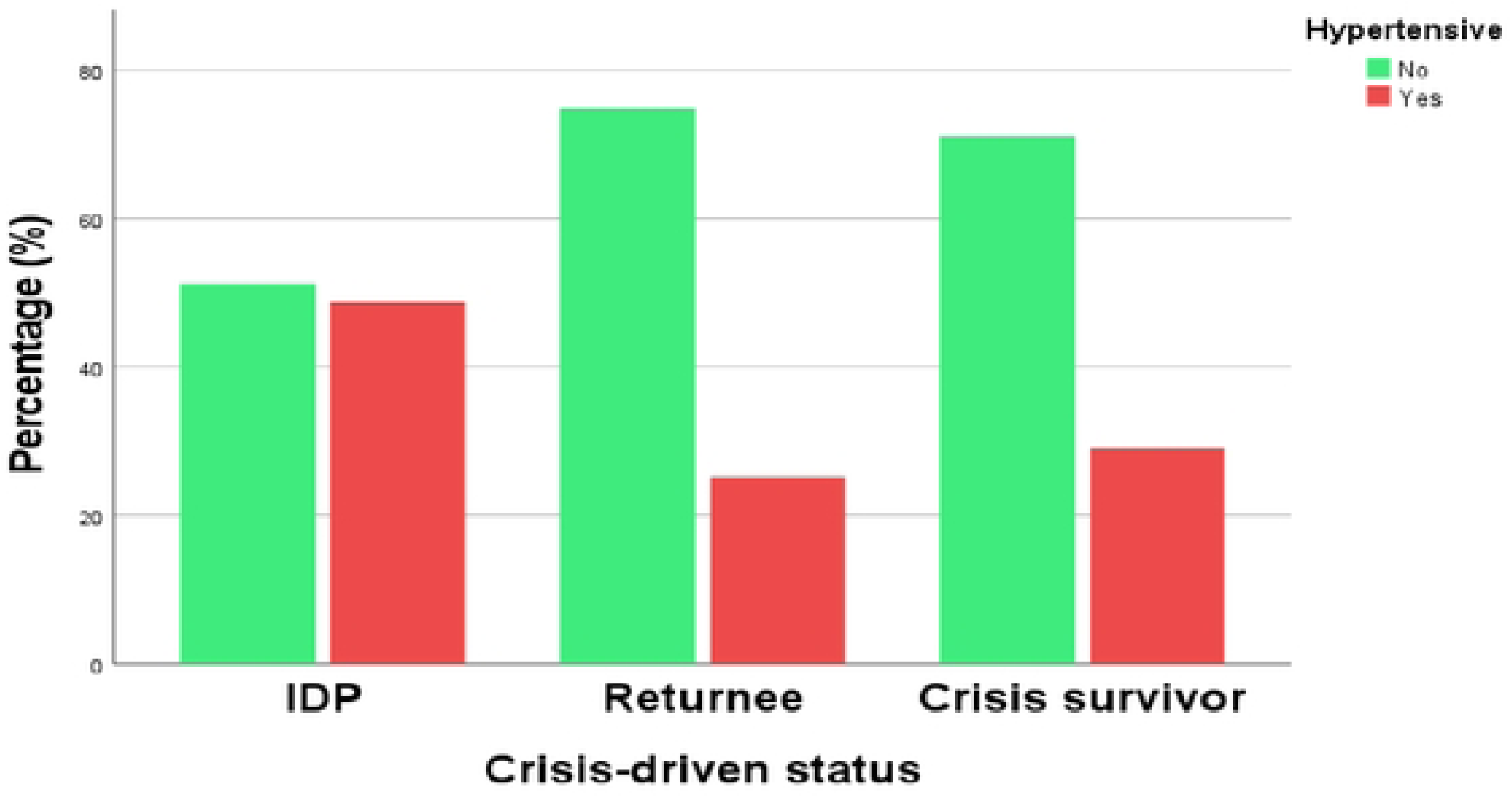
Prevalence of hypertension across crisis-driven status.

Among hypertensive participants (n=264), 63 (23.9%) had controlled blood pressure, 149 (56.4%) had uncontrolled BP and 52 (19.7%) were newly diagnosed cases during the study data collection period.

#### Factors associated with hypertension in conflict-affected areas of Amhara region

In the adjusted analysis of the variables, age was significantly associated with hypertension; study participants in age group 45-54 years and 55-65 years had higher prevalence of hypertension compared to participants in age group 25-34 years (aPR=1.36, 95%CI: 1.03-1.78, p=0.028) and (aPR=1.46, 95%CI: 1.07-2.01, p=0.01) respectively; marital status was also significantly associated with hypertension, unmarried participants had higher prevalence of hypertension compared to those who were married (aPR=1.38, 95%CI: 1.03-1 .94, p=0.033) (Table 2). However, sex, household size, location of residence, educational status, occupational status and wealth index were not significantly associated with hypertension after adjustment.

**Table 2.**
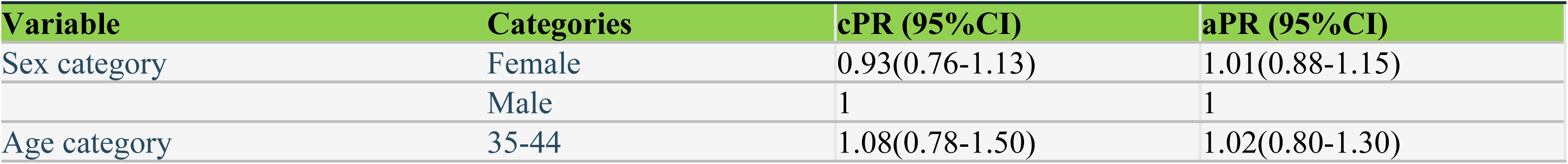
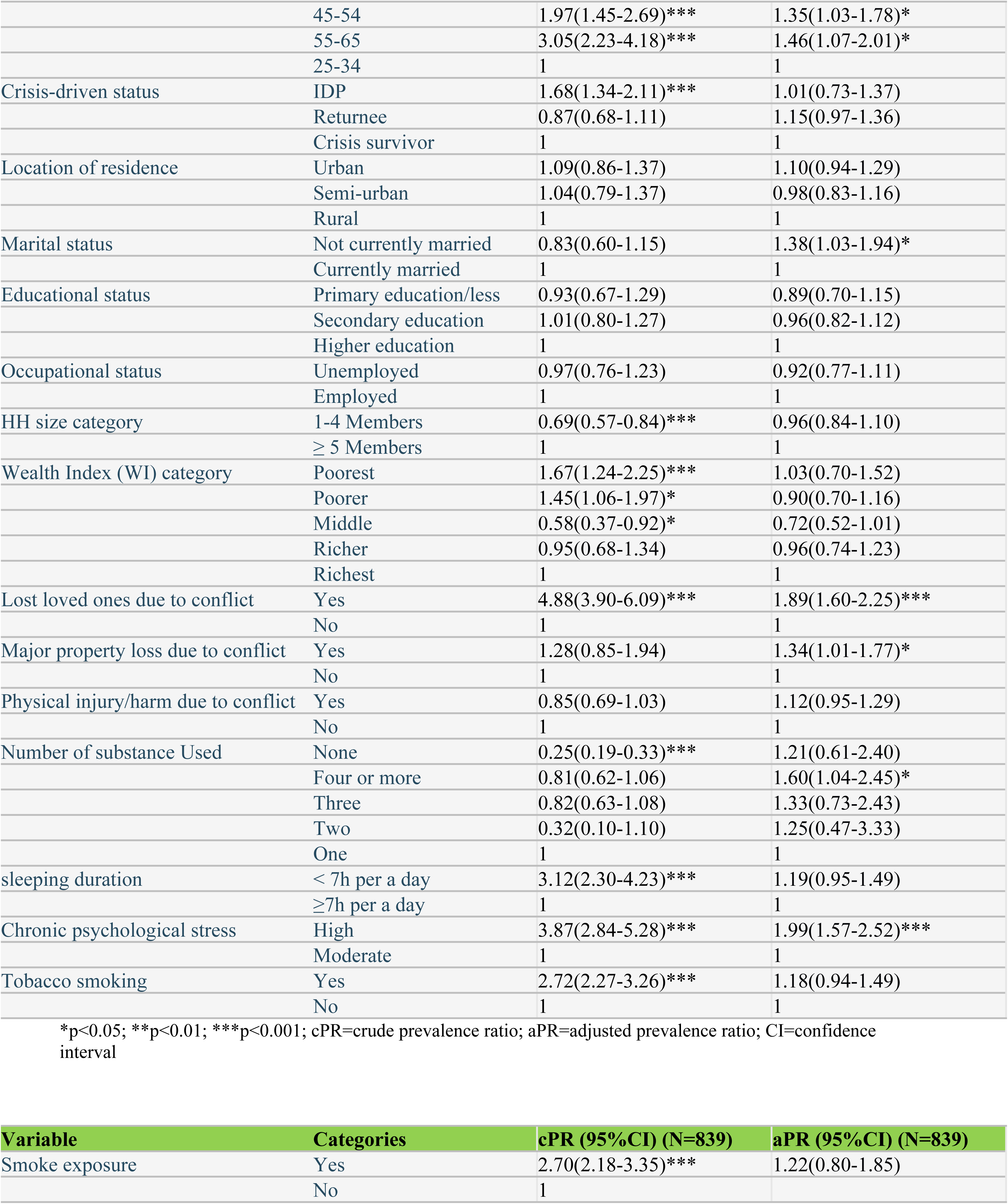
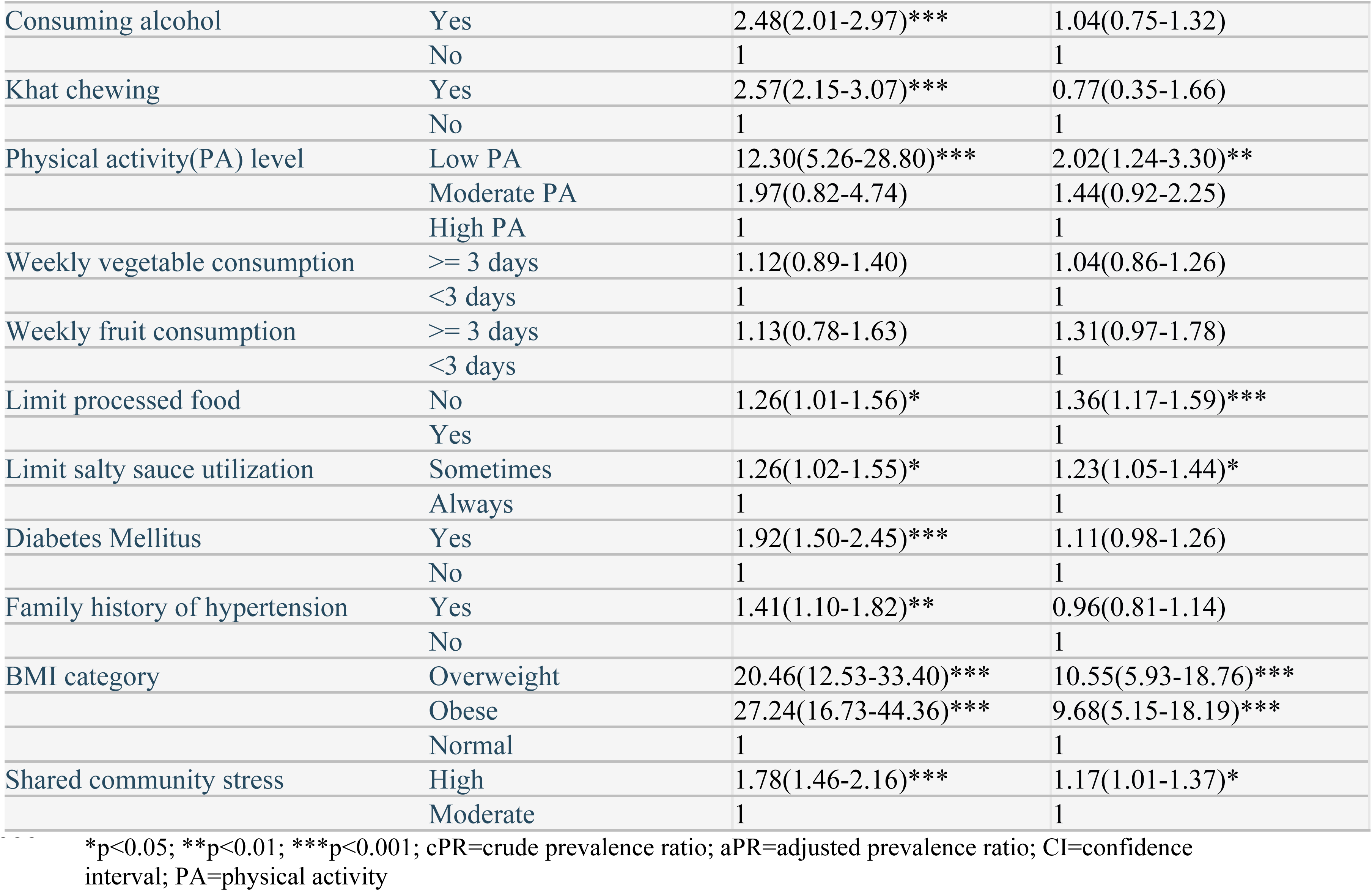
Correlates of hypertension in civil crisis affected areas of Amhara region.

Concerning behavioral factors, study participants with lower physical activity level had higher prevalence of hypertension compared to those with high physical activity level (aPR=2.02, 95CI: 1.24-3.30, p=0.005); participants who did not limit processed for consumption had higher prevalence of hypertension compared to those who limited processed food consumption (aPR=1.36, 95%CI: 1.17-1.59, p<0.001); study participants who eventually limit salty sauce utilization had higher prevalence of hypertension compared to those who always limit salty sauce utilization (aPR=1.23, 95%CI: 1.05-1.44, p=0.012).

Participants who were used multiple substances because of the conflict had higher prevalence of hypertension compared to those used a single substance (aPR=1.60, 95%CI: 1.04-2.45, p=0.034). However, vegetable and fruit consumption, alcohol intake, cigarette smoking, smoking exposure, and khat chewing were not significantly associated with hypertension in the multivariate model.

The psychosocial factors revealed important associations with hypertension; study participants with high level of chronic psychological and shared community stress had higher prevalence of hypertension compared to those with moderate stress level (aPR=1.99, 95%CI: 1.57-2.52, p<0.001) and (aPR=1.17, 95%CI: 1.01-1.37, p=0.043); participants experienced bereavement due to the conflict had higher prevalence of hypertension compared to those did not report such experiences (APR=1.89, 95%CI: 1.60-2.25, p<0.001); participants experienced major property loss due to the conflict had higher prevalence of hypertension compared to those did not encounter such experiences (aPR=1.34, 95%CI: 1.01-1.77, p=0.043). However, variables like suffering physical injury or any harm due to the conflict was not significantly associated with hypertension after adjustment.

Body mass index (BMI) was significantly associated with hypertension. The prevalence of hypertension was higher in overweight and obese participants compared to those with normal BMI (aPR=10.55, 95%CI: 5.93-18.76, p<0.001) and (aPR=9.68, 95%CI: 5.15-18.19, p<0.001) respectively. However, diabetes and family history of hypertension did not remain significantly associated with hypertension in the multivariate analysis.

## Discussion

To our knowledge, this is among the few studies examining biological, behavioural, and psychosocial determinants of hypertension among conflict-affected populations in sub-Saharan Africa. In this cross-sectional study, we found that one in three adults had hypertension and that hypertension tended to be more common in people who were internally displaced.

This community based-study revealed a high prevalence of hypertension among adults living in a conflict-affected community in Ethiopia. The prevalence was higher than the pooled prevalence estimates reported among Ethiopian populations (20.6%) [23] And community-based African populations (27.2%). The high rates of hypertension in our study could possibly be explained by hypertension risk factors such as chronic psychosocial stress, livelihood and health system disruption.

In addition to hypertension prevalence, this study also determined several factors significantly associated with its occurrence: the rate of hypertension increased with age; the finding is consistent with studies conducted in Sub-Saharan Africa countries and beyond [24–25]. This may be explained by age related vascular changes. Our result suggests that hypertension screening and management should be integrated in emergency response health programs. This recommendation is consistent with UNHCR recommendations to integrate NCD services in humanitarian settings [26].

Individuals with low physical activity levels were twice likely to have hypertension compared to those with high activity level. The association between low physical activity level and hypertension is established and explained in several physiological mechanisms; it is associated with obesity, metabolic dysfunctions and establishes other risk factors for hypertension [27]. The result highlights the need to consider physical activity friendly spaces into camp planning and promoting safe, inclusive, and accessible opportunities for exercise in conflict-affected settings.

Processed food consumption was also significantly associated with hypertension. Respondents who did not limit consumption of processed food had higher prevalence of hypertension compared to those who limited processed food consumption. The result is comparable with a study conducted in Iran (one of prolonged conflict countries), which reported increasing incidence of hypertension with processed food consumption [28]. Similarly, other studies conducted in Sub-Saharan Africa links urban diets to high blood pressure [29]. In addition, salty sauce utilization also associated with higher prevalence of hypertension in the current study.

Both processed food and salty sauce utilization can be explained by fluid retention, increased plasma volume and peripheral vascular resistance which contribute to sustained increase of blood pressure due to excessive intake of sodium by individuals [30]. This suggests that the urgent need for strengthened global, regional and national strategies aimed at reducing population-level sodium intake through Policy measures such as food labeling and public awareness campaigns may play a critical role in reducing the risk of hypertension; especially in humanitarian settings.

The current study also revealed that, individuals who used poly-substance following the conflict stress had higher prevalence of hypertension compared to those who used a single substance. This result is consistent with WHO report about substance use and the risk of hypertension, [31], a large synthesis of studies (2014–2024) evidence across Sub-Saharan Africa [32] and a study reported multiple substance use has been linked with cardiovascular strain, poor health and increased likelihood of hypertension [33]. Substance use was common in the studied population suggesting that substance abuse screening and counseling should be integrated in refugee programs, and given attention to reduce its prevalence.

We also found that both chronic psychological and shared/common community stress were significantly associated with hypertension. Respondents with high level stress had higher prevalence of hypertension compared to those with moderate level stress. The finding is similar to global studies exclusively focused on chronic stress as primary exposure and reported independent association with hypertension [34]. In this study three in five adults had stress and stress was associated with a higher risk of hypertension. Stress in conflict-affected settings may be associated to major property loss, bereavement, and displacement; and these were also associated with hypertension. Prior studies [35–37] have reported that stress increases sympathetic drive and therefore stress screening and management should be integrated into humanitarian response programs.

In the adjusted model, Contrary to findings from most studies in sub-Saharan Africa, higher hypertension prevalence was observed among overweight participants than obese ones. This may reflect the unique post-conflict context, where prolonged displacement, food insecurity, nutritional transition, and other contextual factors may have altered the conventional dose-response relationship between BMI and hypertension. Nevertheless, this finding should be interpreted cautiously and confirmed in larger longitudinal studies.

In this study, we also found an association between the established risk factors of hypertension such as overweight and obesity, poor physical activity and ageing. The mechanisms underlying greater hypertension risk for these factors has been previously explained [26–27]. Therefore, lifestyle modification programs focused on diet and physical activity, which could reduce hypertension risk in the displaced population. However, it’s unclear how such life style changes may be implemented in populations facing physical activity limitations due to insecurity and fear.

### Public health implications

Our results have important public health implications. First, health systems should strengthen the integration of hypertension, non-communicable disease, and mental health services through routine community-based screening, psychosocial support, and coordinated models of care that address both individual and structural risk factors. Secondly, district health authorities, healthcare providers, NGOs, and community leaders should collaborate to implement trauma-informed and community-centered interventions that promote healthy diets, physical activity, weight management, and psychosocial well-being while improving access to preventive services. Efforts should also prioritize supportive environments, including safe spaces for physical activity, strengthened community support systems, and livelihood initiatives that can improve health outcomes. In addition, researchers and academic institutions should conduct longitudinal studies for better understanding of causal pathways between psychosocial factors and hypertension in contextual settings.

### Strength and limitations

This may be one of the few studies to document the association between hypertension and structural, psychosocial, behavioral, and biological determinants in conflict-affected populations in SSA. The study identified a broader range of significantly associated factors; for instance socio demographic (age and marital status), behavioral (an emerging factor like processed food consumption), psychosocial (stress, bereavement and major property loss) and biological factor (BMI), which further contribute to policy formulation, immediate and long term public health interventions in a contextualized settings, help address identified research gaps in SSA countries, humanitarian response planning and future research.

Despite its contributions, the study has some limitations; since we measured the exposure and an outcome at the same time (cross-sectional study design), inability to establish causality was the biggest limitation. In addition, internet blackout in some study areas, Selection bias from excluding mentally ill individuals, self-reported nature of many variables and social desirability bias were the concern; however, measures including rigorous training, supervision, and standardized procedures were implemented to minimize potential biases. Therefore, these findings may contribute to the evidence base for hypertension prevention and control in conflict- affected settings

## Conclusion

This study demonstrates a substantial burden of hypertension among conflict–affected populations in Amhara region, affecting nearly one-third of adults, with particularly high prevalence among internally displaced persons. These findings underscore that hypertension in conflict-affected settings is not solely a medical condition but the result of intersecting socio-demographic, behavioral, psychosocial, and biological factors potentially influenced by displacement, disrupted living conditions, and urbanization. The identified factors collectively increase the prevalence of hypertension, highlighting the need for integrated, context-specific interventions. Addressing hypertension in conflict settings therefore requires coordinated strategies that go beyond individual-level care to include social protection, mental health support, and health-system strengthening tailored to displaced and vulnerable populations.

## Data Availability

The deidentified dataset used for the study available upon reasonable request from the corresponding author. Data will be shared subject to approval of a proposal and in accordance with institutional and ethical requirements to protect participant confidentiality. Requests should be sent to

## Acknowledgements

We would like to first express our sincere gratitude to God for granting us the strength, wisdom and perseverance to complete this work. We extend our heartfelt gratitude to Amhara Public Health Institute (APHI), study areas’ zonal and city health department authorities, study participants, data collectors, supervisors and volunteer community health workers. We would also like to appreciate Makerere University School of Public Health, department of community Health and Behavioral Sciences for the overall coordination.

